# Negotiating belonging and identity across the life course: A qualitative study using Social Identity Mapping with people with psychosis

**DOI:** 10.64898/2026.10.27.26365935

**Authors:** Maev Conneely, Veenu Gupta, Janelle M. Jones, Nadia Ayed, Stefan Priebe, Domenico Giacco

## Abstract

A core part of how we understand ourselves comes from the groups we feel we belong to. Having positive, meaningful group memberships, or “social identities”, has been linked to better wellbeing in people diagnosed with psychotic disorders, yet little attention has been paid to how individuals with psychosis experience social identities and changes in belonging across the life course. Personal recovery in psychosis has been described as centred on identity and connection: it may be useful therefore to consider the experiences of people with psychosis from the perspective of the Social Identity Approach to Health (SIAH). This study used Social Identity Mapping (SIM) as the basis for semi-structured interviews (*n* = 28). Thematic analysis, conducted by a team drawing on wide-ranging expertise (lived experience, clinical psychiatry and social psychology), generated four themes: (1) Pre-symptom social identity conflict and non-belonging: *“I’m just something in-between”;* (2) Psychosis tears identity apart: *“The map would have been blank”;* (3) Social identification is both a risk and a requirement for recovery: *“Everyone wants to belong to some certain group or another”*; and (4) Lack of identification with people with psychosis: *“Do I feel a part of that group? Not really, no.”* This SIM-supported interview approach uncovered new insights into how people negotiate social identification before, during and after psychosis. Social identification may be a protective factor, and social identity-strengthening interventions should be evaluated as means of improving quality of life and social functioning.

## 1. Introduction

Social identities, such as being a sister, footballer, or feminist, are central to how we understand ourselves. Social Identity Theory^1^ and Self-Categorisation Theory^2^ propose that a substantial part of our identity, how we understand ourselves, is defined by the groups to which we feel we belong. Applied to health as the Social Identity Approach to Health (SIAH), these theories have shown that maintaining valued group memberships, and developing new ones during major life transitions, is associated with better mental health and quality of life across clinical populations^3–12^. Conversely, disruption or loss of important group memberships can harm wellbeing^12–18^.

Psychotic disorders involve symptoms such as hallucinations, delusions and paranoia, and their onset is often highly disruptive^19^. Qualitative research and first-person accounts suggest psychosis, and treatment, and stigma that can come with illness and diagnosis, can have profound affect on identity^20–26^. However, much of the identity-focused research in psychosis has focused on the patient role and impact of becoming engulfed in the patient role ^27, 28^.

In recent decades, instead of focusing exclusively on the reduction of symptoms, mental health services also aim to support personal recovery for people with psychosis: i.e. supporting individuals to live meaningful, and satisfying lives even with the limitations caused by illness. Personal recovery in psychosis is commonly conceptualised through the CHIME framework, which highlights connectedness and identity as core components^23^. Given the importance of social identification for mental health and the centrality of identity and connectedness to recovery, it is striking that little research has examined how people with psychosis experience belonging, social identification, and changes in these across the life course.

To address this gap, we used the SIAH to understand how people diagnosed with psychotic disorders experience their sense of social identification and changes in their group memberships across the course of their lives. The interview approach was participant-led, aided by visual tool to capture group memberships called Social Identity Mapping (SIM).

## 2. Method

### Design

This was an individual semi-structured interview study reported in line with the Consolidated Criteria for Reporting Qualitative research guidelines (Supplementary Material 1^29^). The study was conducted within a subtle realist worldview with a person-centred axiology^30^. Ethical approval was granted by the Health Research Authority Queen’s Square Research Ethics Committee (18/LO/0799). Participants who took part in a larger quantitative survey (*n*=200) were invited to an interview; 176 agreed (Figure 1), meaning some had previously met the interviewer. Details on recruitment to the quantitative survey can be found in^31^. Guided by lived-experience input from the people in the SUGAR group (Service User and Carer Group Advising on Research, City of London), who had experience of psychosis and/or caring for someone with psychosis, and in line with our worldview, we conducted semi-structured interviews guided by the social identity map that each participant had created as part of the quantitative survey. This supported the aim of exploring how individuals describe their social identities across the life course. SIM anchored discussion in participants’ accounts, and repeatedly brought the interview back to participant-led reflection on how social identities shifted before, during and after psychosis.

**Figure 1.**
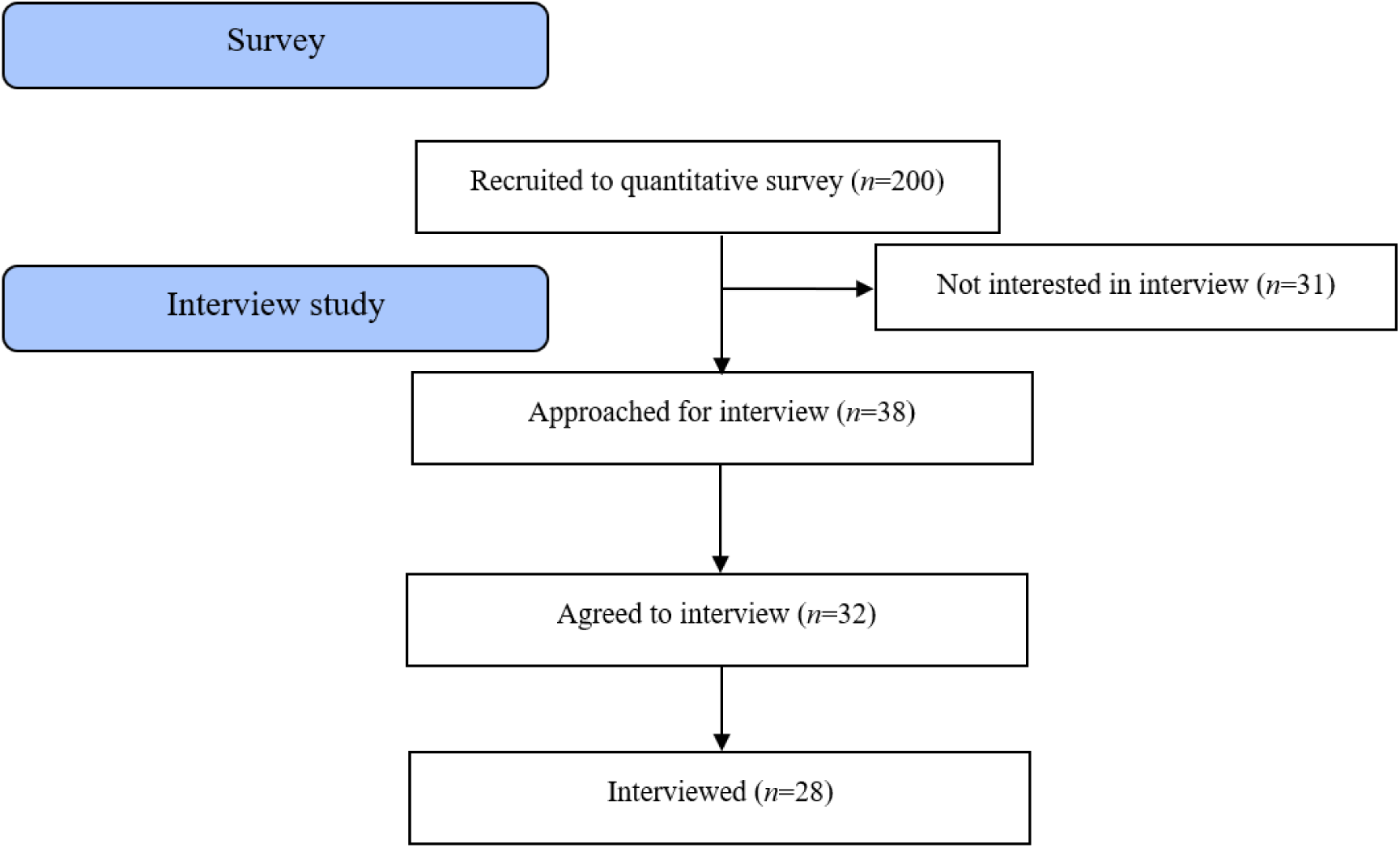
CONSORT flow.

### Procedure

Participants were invited to take part over email or over the phone. Interviews were one-to-one, in private rooms within inpatient and outpatient clinical settings across East London NHS Foundation Trust. All participants were given an information sheet about the study, describing study aims, before deciding to take part and providing informed consent.

#### Topic Guide

A topic guide was developed and piloted by MC, VG (a social identity researcher and expert by experience of psychosis), DG (clinical psychiatrist) and JJ (social psychologist), based on a scoping search and systematic review^28^, and shared with academics in social psychiatry and people with lived experience of psychosis (SUGAR). Following SUGAR’s feedback, the opening and ice-breaker questions were merged into one open question: “People describe themselves in many ways, how would you describe yourself?”. Participants then talked through the map they had created, sharing their thoughts and experiences aloud (topic guide in Supplementary Material 2).^a^

#### Social Identity Mapping

SIM was used to elicit comprehensive data on a person’s sense of social identity, or the ways they see themselves as an “us”^32^. The version used here, adapted for a UK-based population (Figure 2), is described in detail elsewhere^31^ and on the study’s Open Science Framework page (https://osf.io/y867x).

**Figure 2.**
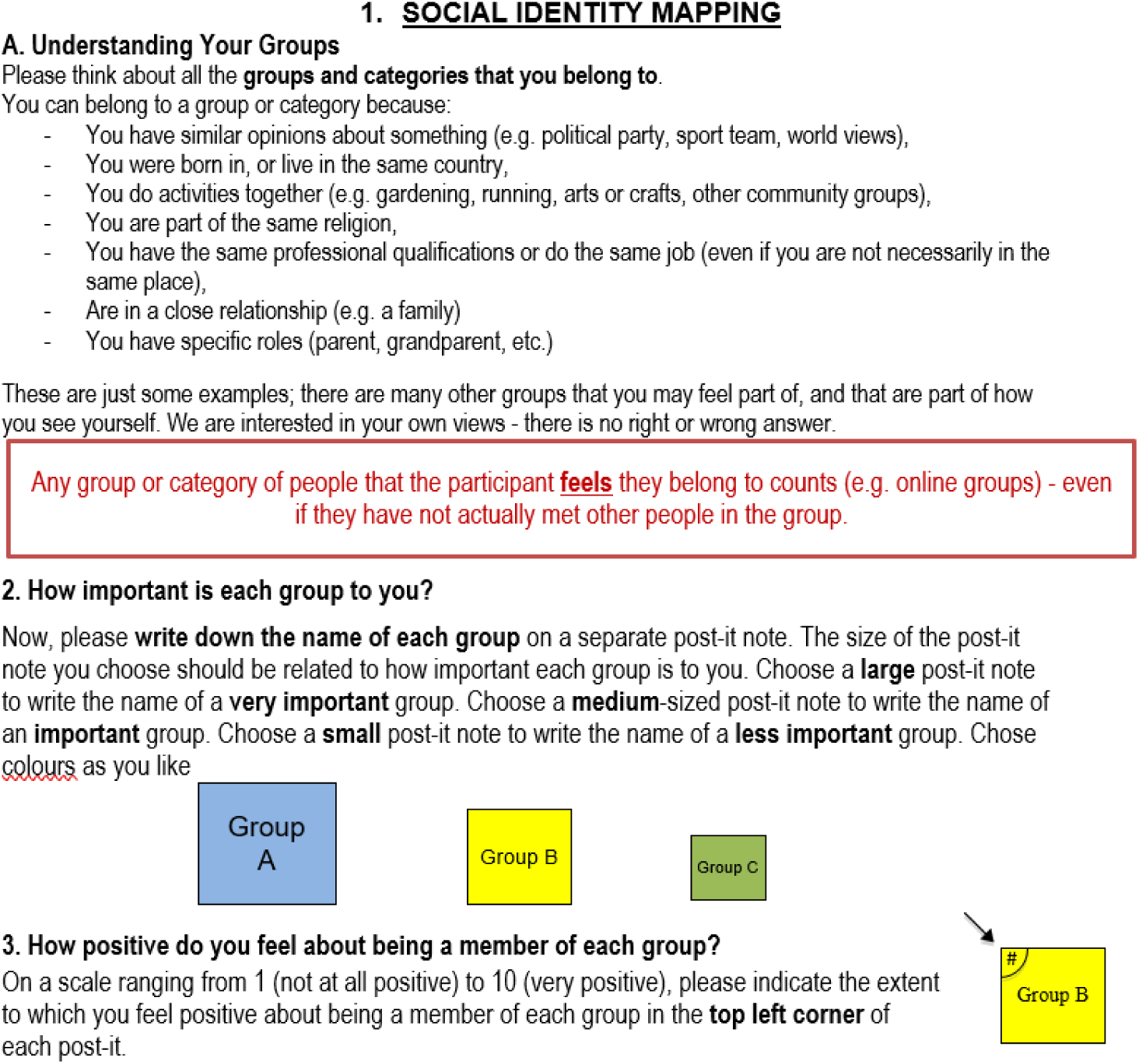
Social Identity Mapping instructions.

### Sample size and purposive sampling criteria

Sampling for qualitative research depends on aims and analysis type^30^. Existing interview research was used to estimate sample size: in a relatively homogenous sample using one-on-one interviews on a single topic, inductive thematic saturation begins at n=12^35, 36^. What constitutes a “homogenous group” here was unclear, so n=12 was used only as a guiding principle. Since number of social identities is a significant determinant of health, wellbeing, and behaviour^4, 5, 8, 37–40^, it was used as the basis for homogeneity. The mean number of social identities in the larger quantitative study this project was nested in was 5 ^31^, so this was used as a cut-off: we aimed to recruit at least 12 participants with more than five social identities and 12 with fewer.

Purposive sampling maximised variation along two axes: 1) time since diagnosis with a psychotic disorder (more or less than 5 years), and 2) number of social identities (above or below the mean of 5). At least n=4 were recruited within each of the four resulting intersections^b^. Time is a salient feature of recovery and identity change^41^, so we sought perspectives spanning 20 years post-diagnosis to the first months of illness. The five-year cut-off operationalised this variation and reflects research suggesting most people with psychosis stabilise around five years after a first episode. Within these categories, we aimed for diversity in gender, age, and ethnicity reflecting the wider sample and population in East London.

Saturation has been criticised as unattainable, since experiences outside the sample will always exist^33^. Here, saturation was understood as a process rather than an event, with recurring ideas indicating shared experiences and reassurance of sufficient recruitment.^34^

### Analysis

Interviews were audio-recorded, transcribed verbatim and anonymised by MC. Thematic analysis followed Braun and Clarke’s approach, combining inductive and deductive coding^42^, which allows for rich, detailed accounts, and provides a useful summary and allows for “unanticipated insights”^43^. Who conducts the analysis matters in qualitative research^44, 45^: in a critical realist paradigm, the analysts’ biases, views and experiences are unavoidable intermediaries in the production of knowledge, so the researcher must be considered part of what is being researched. The analysis team comprised MC, VG, NA, DG, SP and JJ (see above under “topic guide” for areas of expertise). MC led the interviews and analysis (using NVivo) and likely had the greatest influence on the findings. At the time she was a PhD student trained in thematic analysis with degrees in psychology and anthropology (BA) and mental health sciences (MSc). VG, who has personal experience of psychosis and reseach interest in social identity and coded 33% of transcripts, had the next greatest shaping influence in shaping coding and theme-creation.

### Reflexivity

MC, who conducted the interviews and led the analysis, kept a reflexivity journal to consider notes taken during and after interviews. She had previous experience working with involuntary inpatients, and since then had critical views about inpatient mental health services, which could have shaped questioning, tone and analysis. MC is a migrant; which may have increased sensitivity to participants’ accounts of migration, and White, which may have influenced how participants discussed racism and discrimination: interviewer ethnicity affects disclosure of racism, with minoritised interviewees less likely to share with non-minority interviewers^46^.

## 3. Results

### Overview

Twenty-eight people were interviewed. All had a schizophrenia-spectrum diagnosis according to the International Classification of Diseases criteria^47^. Interviews lasted 35-80 minutes. Demographic details are in Table 1; four overarching themes and subthemes are summarised in Table 2. Throughout, participants used the concrete features of the social identity map to explain their sense of who they are, indicating SIM is an intuitive, creative and revelatory experience: *“I thought it was interesting to map it in such a way*” (IDEM181), and, of being in hospital: *“the map would have been blank”* (IDEM181).

**Table 1.** Demographic details Age.

|  |  |
| --- | --- |
| **Age** |  |
|  | 38.14 (*SD*=12.01), range: 19-67 |
| **Gender** |  |
| Male | 15 |
| Female | 13 |
| **Type of treatment receiving at time of interview** |  |
| Inpatient | 13 |
| Community/outpatient | 15 |
| **Detention status under the Mental Health Act (1983)** |  |
| Involuntary | 11 |
| Voluntary | 16 |
| **Ethnic Group** |  |
| Asian/Asian British -Bangladeshi | 7 |
| White British | 6 |
| Black/Black British- Caribbean | 5 |
| Black/Black British- African | 3 |
| White Other | 3 |
| Asian/Asian British- Indian | 1 |
| Other Asian/Asian British | 1 |
| Other Black / Black British Background | 1 |
| Other ethnic group | 1 |
| **Country of birth** | UK (*n*=17) |
|  | Bangladesh (*n*=3) |
|  | Australia (*n*=1) |
|  | Jamaica (*n*=1) |
|  | Latvia (*n*=1) |
|  | Pakistan (*n*=1) |
|  | Poland (*n*=1) |
|  | Romania (*n*=1) |
|  | Somalia (*n*=1) |
|  | Thailand (*n*=1) |

**Table 2.** Overview of themes and subthemes.

|  |  |
| --- | --- |
| Theme 1. Pre-symptom social identity conflict and non-belonging: *“I’m just something in-between”* | 1.1 Stuck without a choice: imposed identities |
|  | 1.2 How you see yourself is not how the world sees you |
|  | 1.3 Tension between different identities leads to uncertainty in who you are: *“Where does that leave me?”* |
| Theme 2. Psychosis tears identity apart: *“The map would have been blank”* | 2.1 Questioning what is real and what can be trusted includes questioning identity |
|  | 2.2 Losing people, roles and activities makes you lose sight of who you are |
|  | 2.3 Changes in thinking processes leads to identity change |
|  | 2.4 Internalising stigma leads to identity change |
|  | 2.5 Identity change is transformation and evolution: *“Killing the person you are, and rise like a phoenix from the ashes”* |
| Theme 3. Social identification is both a risk and a requirement for recovery: *“Everyone wants to belong to some certain group or another”* | 3.1 Isolation is a barrier and a self-protection strategy |
|  | 3.2 Hospital isolates and is proof of disconnection |
|  | 3.3 The route to knowing who you are is through social connections |
|  | 3.4 Groups: social contact, something to do, and they help you feel understood |
|  | 3.5 Feeling a sense of belonging does not require social interaction |
|  | 3.6 Psychosis leads to re-organising social connections |
|  | 3.7 Connection can be found by re-negotiating existing social identities |
| Theme 4. Lack of identification with people with psychosis: *“Do I feel a part of the group? Not really, no”* | 4.1 Disagreement with diagnosis *“I just think it is wrong”* |
|  | 4.2 Concealment due to stigma: *“It is a bit of a secret”* |

#### Theme 1. Pre-symptom social identity conflict and non-belonging: *“I’m just something in-between”*

Participants described a longstanding tension and uncertainty about who they were, predating psychosis. This tension arose from lacking agency or choice over their groups, not being accepted in groups they felt they belonged to, and falling “in-between” groups-between their own feelings and how others perceived them, and between social identities felt as incompatible and internalised into their sense of self. Many described uncertainty, aloneness, lack of belonging, and disconnection. Some also found it hard to describe themselves, unable to answer the opening question: “Sometimes I don’t know who I am. I get lost, confused about who I am” - IDEM192

##### 1.1 Stuck without a choice: imposed identities

Participants described profound disconnection how it stifled agency: demographic or category identities were seen as “paper” identities chosen for them rather than by them, which they could not change even if they wanted to. This lack of choice was a common thread, often relating to groups at high risk of discrimination and marginalisation. Describing the Post-it “Eastern European” on her map, IDEM085 shared this sense of being put into a group:

> *“Yeah, I think that people, once you come to the country, all the time they ask you, ‘to which ethnic group you belong to’, and they categorise you even though they don’t mean to. Even if they have no meanness behind it, they still categorise you”*-IDEM085

IDEM182 expressed the same lack of choice: *“I’m Bangladeshi, but obviously I don’t have a choice in that because I was born into this culture. Obviously.”* The lack of choice was brought up by IDEM080 about the social identity named “Black British”: *“I dunno about that one. I can’t change my identity like sort of thing, and go and live somewhere else. It’s like I’m stuck here. I’m stuck here being British.”*

IDEM159 drew a comparison between two social identities she listed on her map: being female, and being Black. She saw both as not particularly interesting or important to her personally, but that she has been put in these groups by the world and that is what makes them important: *“It just feels, like, I’m female,… and I feel female in the same way I feel Black. I wouldn’t want to change it, but it is who I am. I feel what it means to the world is so much more, and different, from what it means to me.”* IDEM159 contrasted this with identities that she did choose, like being part of a theatre group. When asked about the relative importance of the different groups, she chose one of the imposed, category identities as most important:

> *“I feel like if I had to pick one now, I would probably say Black, even though I want to say theatre. That [pointing to “Black” Post-it] is probably more important. It is not that important to me but it is probably more important to everyone else.”*-IDEM159

People described being in a group that was not chosen by them as distressing. In particular, it was distressing to realise a group they are being categorised in, and did not choose, might lead to discrimination:

> *“In this world I have been put into a category that I feel I have to be in, like for example me being Black, that means I have to listen to this kind of music, or be this kind of way… And then finding out that you might be something [pause] that other people might not consider being that cool.”*-IDEM159

##### 1.2 How you see yourself is not how the world sees you

Disconnection was also expressed as arising from the denial of identities, and awareness of the gap between how people see themselves and how they feel seen by the world. It was distressing and destabilising when others contested their social identities, and led to internalising uncertainty in their own sense of self. This can be seen in IDEM181’s experience of an important social identity being denied: *“So the identity struggle I had was from being in secondary school because people were like ‘oh you are not Jamaican, why are you lying.’”* IDEM181 felt a strong connection to her Nigerian and Jamaican heritage, however, her Jamaican-ness was not accepted throughout her childhood and adolescence because of her appearance, and her name:

> *“Because of my face and my dark skin as well, people would always think ‘this girl is just full African. She is lying about being Jamaican’. But, in fact, I grew up in a single parent household with my mum who is Jamaican and I have more of a Jamaican attitude. My name is [removed], it is so strongly African, people would look at me and say ‘no, this girl is just Yoruba, she is African.’”*-IDEM181

For IDEM200 the chasm between how he feels and how the world sees him is deep: the non-acceptance was reflected in a legal struggle. The denial of his Britishness by the state in his not being given a British passport is a rejection of a belonging that he sees as central to his mental health struggles. Not having a passport prevented him from living the life he wanted to live:

> “I have no memory of Bangladesh, so how can you send me back to a place where I have no memory of? All I remember is being British. That was my situation and that’s really the only thing that is stressing me out. That’s the only reason for why I’m here [in hospital] that’s why I drink, I blaze, that’s the only thing that stresses me out. I couldn’t do anything because of that.”-IDEM200

Not having a British passport stops him feeling British, which has far-reaching practical consequences:

> *“I was aware of it [not having a British passport] because sitting in with the other kids, they get to go to school trips when I have to stay back behind, you understand? And that’s what caused me a lot of depression… im’s weaker do you know what I mean? I feel weaker.”*-IDEM200

For IDEM157, this tension and sense of non-belonging are a source of distress. He described being a minority in the UK as a display of non-belonging, which made him feel unanchored:

> *“To be Asian … to be a minority… In a different country … Has always been something which… I won’t say “troubled” but something I always think about, or something that would always make itself … umm… I don’t know … Not in a sense of racism but… to be different, in a different type of community, to be a minority, to be, to see yourself not blending in, not being part of the whole… yeah … <u>so never, never</u> always feeling that you belong^c^.”*-IDEM157

Belonging to two groups left IDEM157 feeling like he didn’t belong to either:

> *“So I’m neither fully Thai, and neither fully British. So I’m just something that’s in-between. ‘Cause if I move in the Thai community, I’m too British to be Thai. When I’m with British people I’m obviously too Thai to be British. So I’m just something in-between.”-*IDEM157

We can see the effect of having an identity questioned or denied in IDEM122’s description. He said leaving when he was a child under the age of 10 was *“a bit of a wrench really*”, he feels Australian, has an Australian passport, however, he did not include it when completing SIM:

> *“Yeah I mean, describing myself as Australian might not be something that could be understood by people. Because I have been in England for so much more time than I was in Australia”*-IDEM122

This sense of not being allowed, not having the right to feel a sense of belonging appears across different people’s accounts in relation to different social identities. Concern over people questioning a group belonging left people feeling disconnected: uncertain, and in-between.

##### 1.3 Tension between different identities leads to uncertainty in who you are: *“Where does that leave me?”*

People described belonging to an identity they did not choose, as an isolating experience. For example, for IDEM182, the social identity “British Bangladeshi” is one of the 4 Post-its she chose for her map. She described “British Bangladeshi” as something negative because of new conflict related to this social identity since getting married to her Bangladeshi husband:

> *“Being a British Bangladeshi… never used to bother me as much. Basically, because I didn’t have a partner from the actual country. And now that I have married someone from that community in Bangladesh, I feel as though I am not always seeing eye to eye with him… yeah, I just feel a bit sometimes… isolated in my marriage? You know?”-*IDEM182

She explained further that a disconnection in values leads to distress and feeling on the outside, and rejected from the group:

> *“[sigh] It is negative, in the sense that my husband is from Bangladesh and he doesn’t always see things from my point of view. ‘Cause his upbringing is completely different to mine. You know? My in-laws and stuff like that. They don’t really see stuff… they, they can’t help it you know? They can’t see stuff from the way I see it. Because obviously, I grew up here and I’ve been educated here.”*-IDEM182

She feels like she is not accepted as Bangladeshi enough, not seen as part of the community:

> *“That means that me being a Bangladeshi hasn’t affected you… basically you don’t see me as one of your own, to actually help me. So where does that leave me? If you don’t see me as one of your own! “(…) That is what communities are for, you should help each other! Because you know, you are from the same background, you know? But they are not doing that, (…) no, they don’t accept me”.*-IDEM182

Whatever the cause, many participants described feeling unmoored and unsure about their sense of social identity. Often these experiences in conflicts between social identities, and social identity changes were unrelated to psychosis and started before the onset of psychotic symptoms. For example, IDEM033 said:

> *“I had to find my own way (…) I have just been fending for myself my whole life but… I mean, even around my family I don’t feel like I fit in. It is like I am kind of a stranger to myself sometimes. I really don’t know what I want to do.”*-IDEM033

IDEM033 saw this “not fitting in” as having started in childhood - he remembers a television programme about a dog without a home:

> *“A dog in America (…) He just used to travel from place to place, he had no real family or anything. And growing up that is how I used to feel. Like that was what I was gonna turn out like. It just stuck on my mind. No family, and nowhere to call home”-*IDEM033

IDEM033 saw this sense of non-belonging as having originated from his childhood and having been neglected: it appears to have stayed with him.

Participants described internal struggle with social identity, and feeling disconnected from groups, and not having a strong sense of who they are that predates psychotic illness, but which is later compounded by experiencing psychosis and being involved with services. IDEM084 described one of his identities on the SIM as being “*an immigrant*”, even though he was born in the UK and is not an immigrant. The social identity he chose to write on his map tellingly was not the country where his parents were from, but being “an immigrant” - evoking a sense of non-belonging.

#### Theme 2. Psychosis tears identity apart: “*The map would have been blank*”

Psychosis was described as an eraser of social identity and a barrier to forming new ones, often using violent imagery: a force that “tears” identity apart. Many said that after its profound effect on their lives, they no longer knew who they were. That participants saw pre- and post-illness selves as different people was evident in how they responded to the opening question: IDEM185 asked “Now or before the mental health?”, and IDEM144 asked whether to describe herself “as a patient? Or as a person?”. Needing this clarification may reflect being treated differently since becoming a person with psychosis, carrying the assumption that the answer differs, and the implied suggestion that being a patient is somehow not being a person.

The change itself was frequently expressed using violent language:

> *“I’m the same person, of course, the same person torn apart.”-*IDEM064

> *“Umm for my analysis of the - the episode, (2 second pause) it, it just tore my identity apart”*-IDEM157

Many people said that others would describe them as no longer the same person. IDEM080 said it is “*difficult for me to describe myself at this point”*, while IDEM144 said that, while unwell, she is a different person: “*Sometimes when I’m going through that condition, my brother and sister say that I am not myself: ‘You are somebody else’*”. Similarly, IDEM183 expresses uncertainty: *“I don’t know myself as a person.”* IDEM157 explains in more detail the same difficulty:

> *“… that’s a difficult one…ever since after, going through, the episode, my self view … of myself has completely deteriorated. I, I have no idea. I don’t know how to describe myself, or what I am or, who I am. It’s just going with the flow of how things are. I am this, I am this, I am this because a situation determines that I am this. But to see myself^d^ as who I am; not clear.”*-IDEM157

The symptoms of psychosis are seen as barriers to IDEM144 being herself, and leave her unsure about who she is:

> *“Cause yeah… I am a patient. And I do want to become like a normal person. I fight every day; I struggle every day to be one. But I can’t be, because of the hearing voices and things going on with me. Other than that, I am a mother, I am a wife. I have mood swings. I have really low moods. I have really low mood. I really want to be happy in life, but these things, they stop me from being happy, from being myself. It is like I don’t even know who I am at the moment. I don’t have any identity to tell you the truth”*-IDEM144

This erasure of social identity was described in five ways, covered in the subthemes below: making people question their whole reality, including how they saw themselves; losing the activities and connections that anchored their sense of self; a profound change in how they thought, making them feel they could no longer be the same person; internalising the stigma of a devalued group; and, finally, identity change as a distressing but meaningful transformation.

##### 2.1 Questioning what is real and what can be trusted includes questioning identity

Many participants described deeply questioning reality when they were in active psychosis. Questioning everything that they had previously relied on and trusted extended to themselves.

> *“I had an existential crisis. I was questioning whether I existed, whether my friends existed. I went to search their names on the internet and I couldn’t find them and then I was trying to jump out of a window. (…) looking at my friend’s address and seeing nothing coming up and being like what if they don’t exist? What if I don’t exist?”*-IDEM181

IDEM084 sees doubt creeping into who you are as a result of questioning everything else including your senses:

> *“When you get ill… and you have hallucinations and delusions and stuff, it makes you doubt everything. It makes you doubt absolutely everything. You doubt what’s real and what is not. You doubt yourself.”*-IDEM084

##### 2.2 Losing people, roles and activities makes you lose sight of who you are

For many, becoming unwell involved losing social identities, often through the loss of roles and relationships.

> *“Before, there would’ve been far more things I would’ve written down, more than just, just that few there [indicates SIM]”*-IDEM157

People described losing friends through becoming unwell, whether through their own withdrawal or because their friends keep in touch through their admission to hospital: “*my relationship with my friends completely changed. When I became ill, they didn’t respect me anymore really”-*IDEM052

IDEM159 describes not being herself because she was no longer doing the activities she used to do. She listed activities: seeing friends, writing, theatre and then said all became “*crossed out”,* pointing to the SIM.

> “I did feel like performing arts wasn’t important to me, ‘cause I didn’t go to theatre on time, all the time. Or writing-I didn’t do it anymore, or read or anything. Friends. If they didn’t understand me, I didn’t see them. Family-didn’t understand me, didn’t see them. It just became crossed out.”

The loss of the social identities on the map reveals a view of identity is social:

> *“No matter how small or big, these [Post-its] are pillars of who you are. If you start losing them, it is like, you don’t really know who you are.”*-IDEM159

IDEM092 talks about what his SIM would have looked like when he first became unwell, before finding a support group: *“There wouldn’t have been any groups that I identified with. It would have been a blank piece of paper.”* He describes a total lack of group belongings after developing psychosis, using the image of a blank map.

IDEM159 sees the loss of people being replaced by something else: *“I think back then it had to do with a lot of voices. (…) So instead of people, I had the voices in my head*”. She sees the loss of people and the advent of voices as happening hand-in-hand.

##### 2.3 Changes in thinking processes leads to identity change

People felt unable to think in the same way after psychosis. IDEM052 describes his logical reasoning as changed:

> *“My logical intelligence is not good at all, like so I did a first aid course the other day. I can read for hours and understand what I am reading, then two weeks later, I cannot remember a thing. [laughs]. And that is because of my mental illness. I just can’t think the same as I used to, my thinking is changed”*-IDEM052

This change in ability to think is also seen as causing the loss of an aspect of his identity; his sense of spiritual awareness:

> *“When I became ill, I sort of felt like I had lost that aspect of me…My spiritual awareness”*-IDEM052

IDEM107 describes something similar, saying she “thinks slower”: “*I think, I think a bit slower than normal people. (…) I think that people are still the same when they get psychosis, but they think differently because of the diagnosis.”* IDEM157 also sees the way he thinks as a barrier to being the person he was before the episode: “*Yeah ‘cause before the episode, I …. ever since then it’s … the mind just doesn’t work the same way anymore…. and it just doesn’t think.”*

##### 2.4 Internalising stigma leads to identity change

People saw psychosis and being unwell and in touch with services as a failure that meant they had low social status or were “weak”. When asked about the group of people with psychosis, IDEM084 shared:

> *“I don’t talk about it much. I don’t get to talk about it with people who don’t know. So it’s not a big thing, it’s a bit of a secret maybe. Yeah it’s, like, a secret”*-IDEM084

Interviewer: “How does that feel?”

> *“How does that feel, having to have that secret. Em, like I said there’s people’s views of mental health, and they are not very positive in my experience, so it feels … it makes me feel insignificant in a way.”*-IDEM084

IDEM080 describes something similar. When asked if he thought the diagnosis affected how he sees himself he said: “*Sort of, like, my status, like the way, I cannot achieve something. It, sort of, like, blocked it*…*. It makes me look weak*”

The stigma attached to psychosis was internalised for IDEM064 when he was first diagnosed: he describes feeling dirty as a result of being diagnosed: “*I felt dirty. I felt very, very dirty. I felt like I had lost hope in life. I felt like there is nowhere for me in society. I felt that I couldn’t belong anywhere. I felt I am just trash, complete trash, that’s how low I became.”*-IDEM064

##### 2.5 Identity change is transformation and evolution: “Killing the person you are, and rise like a phoenix from the ashes”

The final way participants describe psychosis erasing their old self and bringing transformation. People saw their old self as *“killed”* by psychosis - in order for a new self to be born. IDEM092 describes hearing command voices - and how changing the relationship with the voices is key for many in his hearing voices group:

> *“Most of them hear command voices (…) and the common denominator is ‘kill yourself’. Everyone in the group has had that voice, and is having that voice. But what I say to them is, when you are dealing with the psyche, it is metaphorical and symbolic. And when the voices say ‘kill yourself’, it is like tarot cards. If you get the death card in tarot, it means change. So, if the voice says to you, ‘kill yourself’, they really mean is kill the person you are, and rise like a phoenix from the ashes.”*-IDEM092

For IDEM092, this new person coming out of the ashes is in no way lesser, or worse because of the change, but a *“better person”: “The voices tell you to die. They don’t really want you to die. They just want you to change, become a better person, a more understanding person with more insight.”*

IDEM086 had described a change in herself but also says that she is the same person in a lot of ways – but described change: “But nothing much has changed, I’m still the same person in a lot of ways. Maybe a bit deeper, but you know that’s a part of life”-IDEM086

#### Theme 3. Social identification is both a risk and a requirement for recovery: *“Everyone wants to belong to some certain group or another*”

Threaded throughout participants’ narratives was a need for belonging. Simultaneously, many recognised in themselves a protective reflex to isolate, and withdraw, which was a barrier to experiencing social identification.

##### 3.1 Isolation is a barrier and a self-protection strategy

People described isolation, and the loneliness that comes with it, as intentional. Withdrawing from others was accompanied by a simultaneous longing to belong. For IDEM192, who described herself as not knowing who she is, isolation helos her feel safe in her sense of self:

> *“When I don’t know who I am… staying on my own helps me. I isolate myself. It doesn’t help me in the long term but short term it makes me feel safe, and then I know who I am, to a certain extent”-*IDEM192

We can see IDEM192 was not sure that in the long-term isolating herself helped her, but also says that solitude allowed for more certainty and self-knowledge. Similarly, IDEM157, realised how much he needed people, and how important the connections he does have are to him, while also withdrawing: “*nowadays I’m a hermit [laughs]. I believe somewhere along the line I must’ve chosen to become a hermit”.* He goes on to say he would not choose to be a hermit now: *“Umm, No. no, after going through that experience it, it taught me that I still needed people. I still needed people. I probably wasn’t strong enough to go through that without…well to go through the episode having the support of family. [Family] was really one of the key things to bring me back into normality, the love, the care and the support of the family which then made me appreciate that I needed people*.” - IDEM157

##### 3.2 Hospital isolates and is proof of disconnection

Hospital was the peak of isolation and disconnection, echoing the lack of choice, agency and powerlessness described in Theme 1. Like family, hospital is spoken of as something you do not get to choose:

> *“Them two, you have to be a part of, you have no choice, they force you to come to hospital, and you are born in your family.”* - IDEM193

IDEM200 talks about hospital as a proof of his disconnection, and mirroring his disconnection in the way he feels in his home country, the UK, that is threatening to deport him: *“Yeah, I feel like I’m floating around, I feel like I don’t have a home. Look at me, I’m stuck in hospital.”*

The hospital is described as an in-between, liminal space of non-belonging. For many, diagnosis and hospitalisation after a first episode happened around the same time; together, these confirmed a disconnection many already felt long before symptoms developed, reinforcing otherness and division rather than connection and healing, as IDEM180 describes: “Right now, my life is in stagnation because I am here”

IDEM085 describes the hospital as a lonely place to have been, and talks with gratitude at having been able to get out: *“This is better than being in the isolated place I was in in hospital.”* IDEM193 sees the hospital as a barrier to feeling connected to his British identity: “*If I wasn’t in hospital… I might feel more a part of the country I am living in, not being trapped in hospital, that takes you away from things.”*

This prevented feeling strong ties to social identities:

> *“Truth be told, when I was in hospital, I don’t think was part of anything then. Yeah. It was a tough time, I blocked a lot out. I definitely, definitely wasn’t part of any groups at that time I don’t think”*-IDEM102

The way people talked about hospital was as an in-between place, out of time.

##### 3.3 The route to knowing who you are is through social connections

Social connections, and finding old or new roles are a route out of suffering, and knowing who you are. IDEM159 described how over time the social identities that were lost to psychosis were “built back again”:

> *“It wasn’t as easy, but it happened, so I just had to keep building it back, like I just had to go to theatre like how I normally did. I would start writing like I normally did, I’d see friends like I normally did, saw my family. I’d start doing the things that I loved again and that is what sort of brought me back.”*

Connecting with people again was seen as the way to come back to herself. She directly contrasts reconnecting with social identities by doing the things she used to do, with her admission to hospital, saying the admission was less helpful:

> *“The mental facility for me was almost like telling you how you are meant to act in the world, and then once I was slowly getting out of the mental facility-psychosis was still there but I started doing this, and then this, and then this (pointing at the Post-its), and then understanding what was happening. That is what helped me more.”* - IDEM159

The relationship between how people see themselves and interactions with others is described by IDEM152, and how working on both with psychological services helped:

> *“[Psychological services] definitely affected the way I see myself (…) it helped me create better… it helped me to improve my relationships. [Working with psychology] helped me to improve my self-perception, and when you do that, when you improve that, everything that surrounds you, your relationships, improves as well.”*

IDEM159 saw psychosis as the opposite of connectedness and the removal from the connections as leaving her vulnerable. She describes the process of social identities being ripped away from her. When these social identities that are grounding her sense of self are removed, what was left was psychosis:

> *“It wasn’t like… basically things were being ripped off. So, like if I had to describe the post-its, maybe it was like a part of performing arts got ripped off. And then maybe a part of this was getting ripped off, and all the post-its were getting smaller and smaller until they were just nothing(…) there would just have been one Post-it, just psychosis.”* -IDEAM159

##### 3.4 Groups: social contact, something to do, and they help you feel understood

People describe the benefits of going out, having things to do and a chance to interact with other people. For IDEM192, having a dog is a way of having social interaction with other dog walkers:

> *“’Cause when I walk her, I often talk to people (…) it is just nice to have conversation, ‘cause when you are walking around people don’t want to talk to you on your own, but they will talk to you when you got a dog, won’t they?”*

People talked about mental health support groups as important ways of feeling understood, and having social contact. IDEM055 goes to mental health support groups in the community because *“it keeps me occupied”.* For IDEM144 groups prevent isolation: *“If I do not have a group, I am getting isolated again.”*

Not feeling alone and sharing struggling with mental health was another benefit of mental health support groups, or therapy groups: “*Going to talk about your feelings is really hard, but then you get to hear how they’re feeling and you feel a little bit better. It’s not just me that feels like this, somebody else has this as well, so that really helps”-*IDEM065

IDEM092 says “*All of them have a story to tell and the group is the right place because you have receptive ears. People are like* ‘*oh yeah, I get that’ and it is a problem halved” (…)*

> *“If you say to someone ‘yeah, I understand where you are coming from and reply, then they can cope’, ‘cause they are not on their own.”* - IDEM092

Mental health support groups on people’s maps as social identities were also described as helpful in reaffirming other roles and relationships. For IDEM092 being in a hearing voices group gives “*confidence, self-esteem, the ability to talk about past experiences and learn from the experiences.”* IDEM144 describes going to a mental health support group for Asian women, which helped her because she believed and trusted them. “*Well the Asian women’s group would say that everyone has an identity, they would say you are a mother, a good mother…. [they say] Think positive! I think like that, I think ‘yeah I made sure they [her children] did their homework’”.* IDEM144 says when her kids, or the women at the Asian women’s group, tell her that she has a social identity as a good mother then she can feel it, and she can start to believe it. “*When **they** [members of the Asian womens’ mental health support group] remind me I was a good mother, and I was a good daughter then I can feel that… It is about shared experience*.” IDEM144 listens to what the group tells her because of the shared experiences and values, so she can believe them: “***They*** *understand me, only they understand me.”*

##### 3.5 Feeling a sense of belonging does not require social interaction

Participants also derived belonging from social identities that did not necessarily involve having social interaction. For example, IDEM122 is a bird watcher: the solo nature of the activity does not stop him from feeling a strong sense of connection to, and belonging with, other bird watchers:

> *“When it comes to bird watching I feel like I’m in a fairly large crowd, but they just all do it individually (…) Bird watchers as a group go bird watching on their own. We are all bird watchers, we’re just not all doing it together.”*-IDEM122

IDEM064 also talks about an activity that is usually carried out alone-writing-but shows how connected he feels to other writers, in describing that other writers would agree with him and say the same thing, that they are joined together in their work and in the moments of solitude that the work involves: *“For it was solitary, I like focus, and I think if you interview 10 writers, probably most will say that they have some solitary moments.”* IDEM144 talks about feeling closer to God through prayer. Although it is solitary, and she does not leave home, she feels connected to something bigger:

> *“Yeah, I feel closer to my own creator if I pray. You know praying doesn’t always mean that you sit on a prayer mat and pray. Praying can be in your home, it can be through your heart. I feel closer to my creator.”*-IDEM144

This is also true of IDEM122, speaking about volunteering: he feels a more positive sense of self in that he is part of a greater whole: *“I am, and they are all little cogs in the conservation department.”* His volunteering work to help maintain the canals means he is “peripherally” part of all the conservation going on in the whole country, which helps his mental health.

> *“Eventually it all sort of hooks up into one big conservation act.”-*IDEM122

##### 3.6 Psychosis leads to re-organising social connections

For some people, becoming unwell invited re-evaluating relationships: leading to the loss of some relationships but also to the strengthening of others. IDEM157 shared:

> *“I was close with my father, but not there for my father… and during the episode while I was hospitalised my father would come visit me every day and even though he lives on the other side of London, he would still come visit me every day, and he was really worried and that’s … that created a greater sense of … of bonding with my father”*-IDEM157

Others described similar shifts: “becoming unwell changed my friendships because some people just couldn’t stick around and be there for me, because it was too much for them or it’s made my friendships stronger because they’ve been there for me in the times that I’ve been low” -IDEM065

IDEM052 describes a similar change in his relationship with his family. Before the episode he said he “*wanted to be more independent. I always wanted to leave them rather than join them”* Whereas now he sees his family as a key social identity: *“It is like they are my, my home.”*

##### 3.7 Connection can be found by re-negotiating existing social identities

Identities that were a source of distress previously changed and became sources of support. For IDEM181, previously, her Nigerian name and appearance were seen as barriers to being accepted as Jamaican. However, after an internal process that she went through, of reading about and engaging with her Nigerian social identity, IDEM181 began deriving benefits from this same group she was distressed at felt was imposed on her by others. IDEM181 describes how helpful being accepted unquestioningly as Nigerian became. The same identity, described in Theme 1 as a source of disconnection, became the opposite, an important source of support for her.

> *“Yeah, yeah, no it is not stressful now. Well, it means that African people recognise me straight away as a kindred spirit, as one of them, and it is because of my name and my face. They see that I am Yoruba.”*

The unambiguous belonging and acceptance she now feels contrasts sharply with the distress she felt as a child who was brought up by a Jamaican mother and yet who was not accepted by children and adults at school as Jamaican. She highlights how embracing her African heritage has positively impacted her: *“This Jamaican-Nigerian thing, I really like it right now and it is helping me a lot. When I see the Nigerian staff, they are always friendly towards me because they see my full name and they say ‘this girl is a British Nigerian,’ and then some of the staff, when I was in the hospital would take the mick out of me: ‘The way you speak Yoruba is just so bad!’ [laughs] and I would just be like ‘I don’t understand a word aunty’ [laughs].”*

Others also re-negotiated identities, finding a sense of belonging with those who felt “*in-between”.* IDEM157 described the distress of not feeling fully Thai or British, but finding comfort in meeting others who shared this “in-between” status. He recalls how this commonality brought him and his partner closer:

*“It used to be quite distressing, (…) everyone wants to belong to some certain group or another, but as you grow older with it, it’s just becomes umm… well no there’s, you realise that there’s other people, that are exactly the same way, so you just get on with it, really.”*

#### Theme 4: Lack of identification with people with psychosis: *“Do I feel a part of that group? Not really, no.”*

Most participants did not describe psychosis-related identities, even those who put attending mental health support groups on their maps. When asked if they identified with their diagnosis, most expressed detachment: “*no”* (IDEM086) or *“not really, no”* (IDEM185). They viewed psychosis as an imposed identity, similar to others as described in Theme 1.

##### 4.1 Disagreement with diagnosis: *“I just think it is wrong”*

Participants rejected the diagnosis, some felt it was incorrect. As IDEM122 stated, *“I think the diagnosis is a mistake, so no… I … I just think it is wrong*” Taking the specific diagnostic group out of the question, and asking about the other people who come to the clinic where the interview took place, IDEM122’s response is similar: *“other patients… do I feel part of that?… Not really, no*.” Other people explained that they do not tell their care team that they think the diagnosis is wrong, but that in truth, although they are accepting and following treatment, they do not believe the diagnosis they were given is correct: *“I’ve just gone along with them, but it’s like, I don’t know, I don’t think I have got schizophrenia. I don’t think I have”* (IDEM080).

People who were in hospital, and thus more likely to be interacting with other patients with severe mental health problems, also described not feeling that they belonged to the group of people with psychosis:

> *“There is a lot of different people and they all have different lives, different interests. Most of them do not interact with me, with my interests. So, no, I don’t feel close to them.”-*IDEM180

Showing the distance between herself and others with a diagnosis like hers, and between others with no diagnosis, IDEM144 says *“To me there are two categories. There is a normal category and there is people who are suffering like me, but not exactly as me”,* revealing a sense of separation and isolation.

##### 4.2 Concealment due to stigma: *“It is a bit of a secret”*

Internalised stigma led some to hide their diagnosis. IDEM085 shared, *“There’s a lot of negativity about being a mental health patient, so it’s something you hide…. It is something private”.* Further, for IDEM084, psychosis was seen as an identity that challenged and replaced positive, chosen, hopeful identities.

> *“So for that to replace something positive, like being an aspiring young actor, I think just changed me…. I try to shape my identity on things that, that I like to share, I guess”*

## 4. Discussion

Four overarching themes describe participants’ experiences, which were uncovered by combining Social Identity Mapping with semi-structured interviews. Participants described a disconnection and lack of belonging that pre-dated psychosis, leaving many feeling like “something in-between” long before symptom onset. Psychotic experiences were described as a destructive force, erasing opportunities to interact with others and limiting access to sources of social identification. Belonging was both a risk and a resource: feeling rooted was considered necessary for recovery.

The finding that people felt a lack of belonging, a sense of being “in-between,” unaccepted, and alone *before* symptom onset, sometimes from childhood, is novel and not widely prevalent in the psychosis literature on identity change^28^, and should be examined in quantitative longitudinal research to better understand mechanisms. That not belonging is highly distressing fits with thinking across diagnostic groups on the fundamental “need to belong”, which is theorised as a suicide risk factor in the interpersonal theory of suicide ^48^.

Conflict, identity denial and microaggressions are well-documented sources of distress for minoritised groups^49–51^: 61% of medical students in one US sample reported weekly microaggressions, with a dose-response relationship to depressive symptoms^52^. Memoirs such as Harewood’s (2021) who experience psychosis also describe early racism as profoundly disrupting his social identity, a link emphasised by Oyebode^53, 54^. Given the elevated psychosis risk in minoritised and migrant groups,^55–58^ research is urgently needed into the cumulative impact of identity denial and racist abuse. SIM offered a granular view of how racism and exclusion shape people’s lives; researchers should explore whether social identity contributes to this elevated risk, and whether strengthening social identities alongside policies against discrimination and exclusion could be protective^57–60^.

Psychosis was often experienced as a force that stripped away identity (Theme 2), with hospitalisation perceived as ultimate non-belonging, consistent with prior research^21, 61^. For some, this disconnection was new; for others, it compounded longstanding alienation echoing literature on identity loss through lost roles or stigma, often triggered by symptoms and worsened by hospitalisation^22, 28^.

Belonging, not merely social contact, was considered central to feeling better and living a meaningful life. This echoes the linked larger quantitative study, where number of important social identities predicted quality of life even controlling for symptoms and social circumstances^31^. Together, these findings suggest social identification itself, separate from social contact, may confer benefit. Some participants actively re-negotiated identities through meaningful roles or relationships, which is the aim of the Groups4Health intervention; research should examine whether it could improve clinical and social outcomes in psychosis, building on a promising feasibility study^62^.

Few participants identified with “psychosis” or “voice-hearer” identities. This is notable given the longstanding interest in the “patient”, or illness identity and engulfment of the person into an illness role^27, 28^. Some participants described wanting to conceal or distance themselves from the identity, which is understandable due to the widespread public stigma associated with severe mental illnesses^63–65^ and enduring perceptions that such stigma is legitimate^66–68^.

### Strengths

Strengths of the study include its multidisciplinary team, which alongside working with SUGAR, meant there was continuous lived experience involvement, from the design of the topic guide, through to the coding of the data and write-up. Using SIM to guide interviews helped elicit in-depth experiential data, consistent with recommendations to use visual and creative methods with vulnerable populations to capture more idiographic experiences^69^. The analytic process stayed close to the data, reflected in theme and subtheme names that use participants’ own words rather than prioritising “neat” labels, preserving the complexity and contradictions in participants’ experiences of social identity. The ethnically diverse sample and inclusion of inpatients and people detained under the Mental Health Act ensured representation of perspectives from marginalised gorups, that are often underrepresented or absent in psychosis research.

### Limitations

A first limitation is that all participants were recruited through, and mostly interviewed within, mental health settings (some as involuntary inpatients, others attending outpatient clinics), which could have affected which social identities were salient at that moment (e.g. mental health groups). As all participants were receiving secondary mental health support, the views of those not in contact with services – whether through disengagement, absence of symptoms, or other reasons – are not represented and may differ substantially. Participants who found the initial survey distressing or uninteresting may have declined to be interviewed, though this appears rare (84.5% of the 200 participants of the quantitative study agreed to be contacted); purposive sampling was used to mitigate this risk and ensure diverse perspectives were covered. No repeated interviews were conducted and member-checking was not carried out: as this work considered change over time, the addition of a second interview would have been useful and allowed to uncover the stability over time of SIM. Doing member checking, and reimbursing participants for their time, would have fit better with the axiology of the study. This practice, though not all paradigms or approaches see it as essential, can enhance trustworthiness and credibility^70, 71^ however, due to resource limitations (financial and temporal) this was not possible.

### Conclusion

Using SIM generated insights not previously observed in the psychosis literature on social identity and identity change, particularly, disconnection experienced independently of, and long before, prodromal or active psychotic symptoms. Further research should explore whether social or clinical interventions incorporating social identity-building benefit people with psychosis. Such interventions may address what appeared to be a common experience among participants: contact with services often compounded disconnection rather than fostering connection. Participants lived with a tension around social connection: social withdrawal is considered self-protective, yet it reduces the sources and opportunities for deriving social identification: “*without those pillars…you don’t know who you are*”. More research is needed to understand how services and third-sector organisations can help harness the benefits of meaningful group belonging, in a way that is not overwhelming, and how discrimination and identity denial may shape belonging and, in turn, vulnerability to psychosis.

## Data Availability

Data are available upom reasonable request.

## Supplementary Material

Supplementary Material 1. Consolidated criteria for reporting qualitative research (COREQ) checklist

Supplementary Material 2. Topic Guide

## Footnotes

a Throughout the interview the language used for distress (e.g. “mental health problems”, “psychosis”, “schizophrenia”, “patient/service user”, “the episode”) was taken from the participant, and thus it is inconsistent throughout the excerpts below.

b i.e. diagnosed more than five years with fewer than five identities, diagnosed more than five years with more than five identities, diagnosed less than five years with fewer than five identities, and diagnosed less than five years with more than five identities.

c Emphasis added

d Emphasis in interview

